# Retinal Vasculature, Plasma Metabolites and Cardiovascular Disease Risk: A Prospective Cohort Study from the Canadian Longitudinal Study on Aging

**DOI:** 10.64898/2026.05.06.26352609

**Authors:** Simona I. Indzhova, Philip Britz-McKibbin, Talha Rafiq, Divya Joshi, Fahmida Mannan, Emanuele Trucco, Sandi M. Azab, Michael Chong, Marie Pigeyre

## Abstract

**Background:** This study aimed to investigate whether retinal fractal dimension (D_f_), a measure of microvascular branching complexity from fundus images, together with plasma metabolites, can help identify pathways linking microvascular changes to cardiovascular disease (CVD) events.

**Methods:** We analyzed longitudinal data from a subset of 4,781 participants from the Comprehensive cohort of the Canadian Longitudinal Study on Aging (CLSA), free of CVD at baseline (mean age 58.74 ± 8.39; 47% male), and with 811 plasma metabolites measured at baseline and retinal imaging. Total, arterial and venular D_f_ were derived from fundus photographs using the Vessel Assessment and Measurement Platform for Images of the Retina. Incident CVD was defined as one or more of self-reported physician-diagnosed myocardial infarction, angina, coronary heart disease, stroke, transient ischemic attack, or peripheral vascular disease during follow-up. Regression models tested associations among D_f_, plasma metabolites and incident CVD.

**Results:** Over a median follow-up of 5.75 years (IQR 5.48-6.04), 546 participants (11.42%) developed CVD. Higher total, arterial and venular D_f_ were associated with lower CVD risk in unadjusted analyses (Odds Ratio (OR)=0.62, 95% CI:0.53-0.72 for total D_f_; OR=0.78, 95% CI:0.70-0.86 for arterial D_f_; OR=0.74, 95% CI:0.67-0.82 for venular D_f_). Total D_f_ demonstrated the strongest predictive value for incident CVD but not independently of established CVD risk factors. Nine plasma metabolites, including amino acids, lipids, and xenobiotics, were associated with both incident CVD and one D_f_ measure (p < 0.05), with cotinine and hydroxycotinine satisfying a false discovery rate adjustment (q < 0.05). Consistent with these findings the Total Nicotine Equivalent (TNE-3) was also associated with both lower arterial D_f_ and increased CVD risk.

**Conclusions:** Retinal microvascular complexity is associated with incident CVD. Nicotine metabolism from tobacco smoking exposures emerged as the strongest association linking microvascular changes and CVD events.

**Clinical Perspective:** *What is New?:*

- Lower retinal branching complexity, as measured through D_f_, is significantly associated with incident cardiovascular disease (CVD) in unadjusted analyses.
- In a population-based sample, we found nine plasma metabolites associated with both retinal vascular complexity and incident CVD, but only nicotine metabolites were significant after multiple testing correction.
- Nicotine metabolites, particularly cotinine and hydroxycotinine, remained significantly associated with incident CVD even after adjustment for self-reported smoking status.

*What Are the Clinical Implications?:*

- Retinal D_f_ measures could be used as a predictor of CVD event, although not independently of age and traditional cardiovascular risk factors.
- Microvascular changes may lie on the pathway linking nicotine metabolites to CVD events.
- Plasma nicotine metabolites may provide additional cardiovascular risk information beyond self-reported smoking, reflecting individual exposure, metabolism and passive smoke exposure.

## Introduction

Cardiovascular diseases (CVD), including stroke, myocardial infarction (MI), and peripheral vascular disease (PVD), remain a leading cause of mortality and disability worldwide among older adults^1^. CVD develops through early, cumulative vascular and metabolic disturbances that cause structural and functional microvascular alterations long before clinical symptoms emerge.

The retinal vasculature provides a uniquely accessible window into the systemic circulation. Sharing physiological, structural and anatomical features with coronary vessels^2^, retinal vessels can serve as surrogate markers of coronary microvascular health^3^, a concept already integrated into clinical practice through retinal screening for diabetes and hypertension. Fractal dimension (D_f_), computed from retinal photographs using automated deep learning, quantifies vascular density and branching complexity^4^. Higher D_f_ indicates a more intricate branching network^5^, and lower D_f_ has been associated with increased risk of stroke, fatal stroke, coronary heart disease (CHD), and cerebrovascular events^6^. However, the clinical utility of retinal D_f_ remains uncertain, as it is unclear whether it independently predicts CVD beyond traditional risk factors or primarily reflects underlying cardiometabolic burden.

Metabolomic signatures associated with retinal vascular complexity may provide insights into pathways linking microvascular structural changes that precede CVD. Several metabolites have been linked to increased CVD risk, including branched-chain amino acids, aromatic amino acids^7^, gut microbiota-derived metabolites such as phenylacetylglutamine^8^ and atherogenic trimethylamine N-oxide (TMAO)^9^, as well as lipid-based metabolites, such as lysophophatidylcholines and sphingomyelins^7^. Yet, little is known about how circulating metabolite phenotypes influence the microvascular architecture as captured by retinal D_f_, or whether shared metabolic pathways underlie associations between retinal vascular complexity and incident CVD.

To date, few population-based studies have examined the intersection of retinal microvascular complexity, circulating metabolites, and incident CVD^6^. In addition, although total, arterial, and venular D_f_ can be derived separately, the extent to which vessel-specific measures provide distinct predictive information remains unclear.

Using data from the Canadian Longitudinal Study on Aging (CLSA), we examined the demographic, socioeconomic, clinical, and lifestyle determinants of total, arterial, and venular fractal dimension (D_f_). We then evaluated the associations of circulating plasma metabolites and retinal D_f_ with incident cardiovascular disease (CVD) and estimated the extent to which retinal D_f_ mediates the association between plasma metabolites and CVD risk. We hypothesized that lower retinal D_f_ would be associated with a higher risk of incident CVD events, and that certain plasma metabolites would be associated with both retinal D_f_ and CVD. This integrated approach allows for the identification of metabolic pathways underlying microvascular alterations and provides insights into the biological mechanisms linking retinal D_f_ with CVD risk.

## METHODS

### Data Source and Study Participants

Details of the CLSA including the exclusion criteria have been described previously^10^. Briefly, the CLSA recruited 51,338 community-dwelling adults aged 45-85 years at baseline (between 2011 and 2015) using stratified random sampling to create a nationally representative cohort. Follow-up occurs every three years, with minimal loss to follow-up, as indicated by retention rates of 92% (n = 27,765) in the first wave (2015-2018) and 85% (n = 25,563) in the second wave (2018-2021).

The cohort comprises two components: the Tracking cohort (n = 21,241) and the Comprehensive cohort (n = 30,097). The Tracking cohort includes participants from all 10 provinces who provided data through a computer-assisted telephone interview^10^. The Comprehensive cohort consists of participants who provided data through in-home interviews and attended one of 11 data collection sites (DCS), where they underwent detailed assessments including physical examinations and biospecimen collection. Blood samples were collected from 23,492 Comprehensive cohort participants at baseline. A stratified random subset of 10,000 selected by age group, sex, and DCS was chosen for plasma metabolomic profiling, and metabolomic data are available for 9,992 participants^11^. The study size was determined by the number of Comprehensive cohort participants with moderate or good quality retinal images, available plasma metabolomic data at baseline, available clinical data across baseline, first, and second follow-ups, and free of CVD at baseline (n = 4,781). Ethical approval was obtained from the Hamilton Integrated Research Ethics Board (Project ID: 2304013), and all participants provided written informed consent.

### Retinal Photography and Fractal Dimension

At baseline, 57,434 retinal images were collected from 25,678 participants in the Comprehensive cohort. Retinal photographs were captured using a Topcon TRC-NW8 nonmydriatic retinal camera at a 45° angle, beginning with the right eye. Detailed image acquisition procedures and contraindications have been previously explained elsewhere^12^.

Retinal vascular features were quantified using the Vessel Assessment and Measurement Platform for Images of the Retina (VAMPIRE), version 3.1 (University of Edinburgh and University of Dundee, UK), which automatically identifies retinal landmarks and computes vascular metrics^13^. For fractal analysis, macula-centered images underwent automated vessel segmentation to generate binary vascular maps, followed by manual review to correct artifacts. Skeletonized maps were used to calculate total, arterial and venular fractal dimension (D_f_) using the generalized sandbox method^13^. Briefly, a grid of increasing box sizes (powers of 2) was superimposed over each skeletonized image, and the number of boxes containing at least one vascular pixel was counted^14^. D_f_ was derived from the slope of the linear regression between the natural logarithms of box counts and box sizes. Fundus image quality was automatically graded as *good*, *moderate* or *poor*; only participants with at least one eye graded *moderate* or *good* were included. When both eyes were of comparable quality, mean D_f_ values were used; otherwise, the higher-quality image was selected. All D_f_ measures were quantile-normalized before analysis.

### Plasma Metabolomics

Untargeted metabolomic profiling of non-fasting baseline plasma samples^11^ was performed by Metabolon, Inc. (Durham, NC, USA), using a Waters ACQUITY ultra-performance liquid chromatography (UPLC) coupled to a Thermo Scientific Q-Exactive Orbitrap high resolution mass spectrometer (MS) system interfaced with a heated electrospray ionization (HESI-II) source^11^. Samples were analyzed across 99 analytical batches (∼100 samples per batch) and were randomized both within and across batches to reduce bias. Each sample was divided into four aliquots and analyzed under different chromatographic platforms and ionization modes to expand metabolome coverage. There were two positive ionization modes, one optimized for more hydrophilic compounds and the other for more hydrophobic compounds, and two negative ionization modes, including one on a reversed-phase C18 column with basic elution conditions and one on a hydrophilic interaction liquid chromatography column for polar metabolites. Compound identification was achieved through comparison with Metabolon’s proprietary database, based on retention time, mass-to-charge ratio (*m/z*), and MS/MS spectra acquired with data-dependent MS^n^ scans using dynamic exclusion^11^.

Quality control procedures have been described in detail previously^11^. Briefly, 1,314 metabolites spanning multiple biochemical classes were identified, and metabolites were retained if present in >50% of samples with adequate technical precision. Batch correction followed Metabolon’s standard protocol, whereby raw metabolite values were median-scaled within each batch (setting batch and metabolite medians to one) and subsequently normalized to batch-specific QC samples^11^. Values exceeding ± 3 standard deviations (SDs) were winsorized. Plasma metabolites were then natural log-transformed to reduce skewness and standardized to a mean of 0 and SD of 1, consistent with prior CLSA plasma metabolite preprocessing^15^. A total of 811 plasma metabolites, including 130 of unknown identity, that were consistently measured across all analytical batches, were included in the analyses (Supplementary Table 1). As Metabolon quantifies metabolites as relative rather than absolute concentrations, all analyses are based on these relative values.

### Covariates

Baseline covariates included demographic, socioeconomic, clinical, and lifestyle variables selected a priori. Demographic variables included age (years), biological sex at birth (male/female), and genetic ethnicity (European/non-European). Socioeconomic status (SES) was based on self-reported household income (<$20,000; $20,000-49,999; $50,000-99,999; $100,000-149,999; ≥$150,000), highest level of education (less than secondary, secondary without post-secondary, some post-secondary, or post-secondary degree/diploma), and employment/retirement status (fully retired, partially retired, employed and not retired, never employed, not currently employed but previously employed)^16^. A composite SES variable (low, medium, high) was derived via latent class analysis using complete cases for SES indicators. Anthropometric measures included body mass index (BMI, kg/m²) and waist-to-hip ratio (waist circumference/hip circumference, WHR). Systolic and diastolic blood pressure (BP) was calculated as the average of five out of six consecutive readings (excluding the first) (mmHg). Pre-existing diabetes was defined using self-report of physician-diagnosed diabetes, HbA1c ≥ 6.5% or diabetes medication status (both oral and insulin). Pre-existing hypertension was defined as systolic BP ≥ 140 mmHg or diastolic BP ≥ 90 mmHg, with adjustments for antihypertensive medication use (+15 mmHg systolic, +10 mmHg diastolic)^17^ and/or antihypertensive medication use. Pre-existing dyslipidemia was defined as low-density lipoprotein cholesterol (LDL) > 4.5 mmol/L, or use of lipid-lowering medication, with LDL adjusted for statin use (LDL/0.8)^18,19^.

Lifestyle factors included alcohol consumption (frequency in the past 12 months), smoking status (never vs former vs current), diet quality, and physical activity. Smoking intensity was defined in current smokers using self-reported cigarettes per day. Reported categories (1-5, 6-10, 11-15, 16-20, 21-25, and ≥ 26 cigarettes/day) were converted to continuous values using category midpoints. For participants reporting ≥ 26 cigarettes/day, exact values were used^20^. Diet quality was assessed using the Mediterranean Diet Score (MDS) based on data from the validated Short Diet Questionnaire (SDQ)^21,22^. Dietary components were classified as “beneficial” (non-refined cereals, fruits, vegetables, legumes and nuts, potatoes, fish) or “detrimental” (meat and meat products, poultry, full-fat dairy products)^23^. For beneficial components, participants at or above the sex-specific median received 1 point; those below received 0. For detrimental components, participants below the sex-specific median received 1 point; those at or above received 0^24^. Alcohol consumption was scored separately: moderate consumption (> 0 and ≤ 7 drinks/week for females; > 0 and ≤ 14 drinks/week for males) received 1 point, while 0 or higher-than-moderate consumption received 0 points^25^. Component scores were summed to yield a total MDS ranging from 0 (lowest adherence) to 10 (highest adherence). Physical activity was assessed using a modified Physical Activity Scale for the Elderly (PASE), capturing weekly frequency and duration of walking, light/moderate/vigorous sports, and strength training. Weekly hours of activity were estimated by calculating midpoints for reported frequency and duration, with maximum duration capped at 4 hours to reduce overestimation^26^. Total activity was expressed in metabolic equivalent of task (MET) minutes per week and categorized as low (<600 MET-min/week), moderate (600-3000 MET-min/week) or high (>3000 MET-min/week)^27^.

### Cardiovascular disease events

CVD event was based on participant self-report of physician-diagnosed conditions, including CHD, angina, PVD, myocardial infarction (MI), stroke/cerebrovascular accident (CVA), or transient ischemic attack (TIA). Prevalent CVD was assessed at baseline, while incident CVD was assessed at follow-up 1 and follow-up 2. Participants free of CVD at baseline with data available for at least one follow-up visit were classified as incident cases if CVD was reported at any available follow-up. CVD severity was categorized into four groups based on the number of events: No CVD, mild (1 event), moderate (2 events), and severe (3+ events).

### Statistical Analysis

All analyses were conducted in the full cohort of 4,781 participants, unless otherwise specified. Missing covariate or outcome data were addressed using a complete-case approach. Random forest regression models were first used to identify demographic, socioeconomic, clinical and lifestyle predictors of total, arterial and venular D_f_ among 17 candidate variables, using a 70/30 training-testing split. Model performance was evaluated using R^2^ and root mean square error (RMSE). Variable importance was assessed using the mean decrease in accuracy (MDA) with a threshold of ≥ 20 used to identify influential predictors.

Next, logistic regression with forward stepwise selection based on Akaike Information Criterion (AIC) minimization was used to identify significant predictors of incident CVD among 17 candidate variables. Analyses were restricted to participants with complete data for all covariates and retinal D_f_ measures, as well as those without baseline CVD and with available incident CVD status. Covariates retained in the final model were ranked according to their Wald χ2 statistics (β/Standard Error)^2^. Collinearity was assessed using the variance inflation factor (VIF) > 5.

Associations between each D_f_ measure and incident CVD were examined using multivariable binary logistic regression. Three models were constructed: Model A (unadjusted), Model B (adjusted for age and sex), and Model C (adjusted for covariates retained from the forward stepwise selection). In addition, ordinal logistic regression was used to evaluate associations between retinal D_f_ measures and CVD severity. Linear regression models, adjusted for age and sex, were used to assess associations between total, arterial and venular D_f_ (dependent variable) and each plasma metabolite (independent variable). False discovery rate (FDR) correction for multiple testing was applied to the p-values from each regression analysis (811 metabolites tested per retinal D_f_ model), with statistical significance defined as q < 0.05. Using multivariable binary logistic regression, each metabolite was tested separately for its association with incident CVD, first adjusted for age and sex, and in a subsequent model adjusted for the covariates retained from the forward stepwise model. Associations were expressed per 1 SD increase of natural log-transformed standardized plasma metabolite. False discovery rate (FDR) correction for multiple testing was applied to the p-values from each regression analysis (811 metabolites tested), with statistical significance defined as q < 0.05. To further characterize the observed associations for composite incident CVD, secondary event-specific analyses were conducted for cardiovascular events (CHD, angina, MI), cerebrovascular events (CVA or TIA), and PVD. Logistic regression models were fitted separately for each outcome using the subset of plasma metabolites previously associated with both D_f_ measures and incident CVD. Sex-specific effect modification was evaluated across the models examining (i) retinal D_f_ and incident CVD, (ii) retinal D_f_ and plasma metabolites, and (iii) plasma metabolites and incident CVD.

Finally, mediation analysis was conducted to assess whether retinal D_f_ measures mediated associations between circulating plasma metabolites and incident CVD. Metabolites included were those previously associated with both retinal D_f_ and incident CVD. For each plasma metabolite, a two-model approach was implemented. First, a linear regression model was fitted with retinal D_f_ as the continuous mediator and the plasma metabolite as the exposure, adjusting for age, BMI, pre-existing hypertension, smoking status, WHR, physical activity, ethnicity, alcohol consumption, pre-existing dyslipidemia and LDL cholesterol. Second, a logistic regression model was fitted with incident CVD as the binary outcome, including both the plasma metabolite and the corresponding D_f_ measure, with adjustment for the same covariates. Natural direct and indirect effects were estimated using nonparametric bootstrapping with 5,000 simulations.

All statistical analyses were performed using R Statistical Software (v4.5.2)^28^, using packages such as poLCA, ggnewscale, RColorBrewer, grid, mediation, e1071, patchwork, car, caret, and randomForest. Statistical significance was defined as a nominal (unadjusted for multiple testing) p < 0.05 or q < 0.05 after FDR correction. Nominal p values were initially used to identify metabolites associated with retinal D_f_ measures and with incident CVD; these metabolites were subsequently subjected to FDR correction. All models were specified a priori.

## RESULTS

### Retinal Image Quality and Associations with Ocular Conditions and Retinal D_f_ Measures

Of the 25,678 participants with retinal images available, 25,368 (98.8%) had at least one gradable image (poor, moderate, good) in at least one eye, as assessed by VAMPIRE. The remaining participants were missing image quality grading in both eyes (n = 310). Poor-quality images in both eyes, or a combination of one poor-quality and one ungradable eye, were associated with a higher prevalence of ocular conditions, including cataracts, glaucoma, macular degeneration, and diabetic retinopathy (Chi-square p < 0.0001) (Supplementary Table 2). Retinal D_f_ measures were positively correlated with the image quality (moderate or good) used to calculate them with Spearman correlation coefficients of ρ = 0.65 for total D_f_, ρ = 0.53 for arterial D_f_ and ρ = 0.48 for venular D_f_ (all p < 0.0001, Supplementary Figure 1). After restricting to participants with non-missing incident CVD status (n = 18,165), those with poor-quality retinal images were generally older and had less favorable cardiometabolic profiles, including higher systolic blood pressure, HbA1c, and pre-existing hypertension (Supplementary Table 3).

### Determinants of Retinal D_f_

Further restricting to participants with complete data for all three retinal D_f_ measures and incident CVD status (n = 13,128), among the 17 demographic, socioeconomic, clinical and lifestyle factors examined, age, systolic BP, diastolic BP, sex, and WHR were the strongest predictors of total D_f_ (Figure 1). Age was also the strongest predictor for arterial D_f_, followed by systolic BP, diastolic BP, pre-existing hypertension, and WHR. Similarly, for venular D_f_, age remained the most influential predictor, followed by sex, systolic BP, WHR, ethnicity and diastolic BP. Overall, the determinants of total, arterial, and venular D_f_ largely overlapped, differing primarily in relative ranking across the measures (Supplementary Figure 2). Males had higher retinal vascular complexity than females, with mean total D_f_ and venular D_f_ values that were 0.11 units (95% CI 0.07-0.14, p < 0.0001) and 0.19 units higher (95% CI 0.14-0.24, p < 0.0001), respectively. No significant sex differences were observed for arterial D_f_. Arterial and venular D_f_ were shown to be moderately correlated (Pearson’s r = 0.47, p < 0.0001), whereas total D_f_ showed strong correlations with both arterial D_f_ (r = 0.69, p < 0.0001) and venular D_f_ (r = 0.68, p < 0.0001). Steiger’s test for dependent correlations indicated no significant difference between the two correlations (p = 0.10), suggesting that total D_f_ is equally strongly associated with both arterial and venular D_f_.

**Figure 1.**
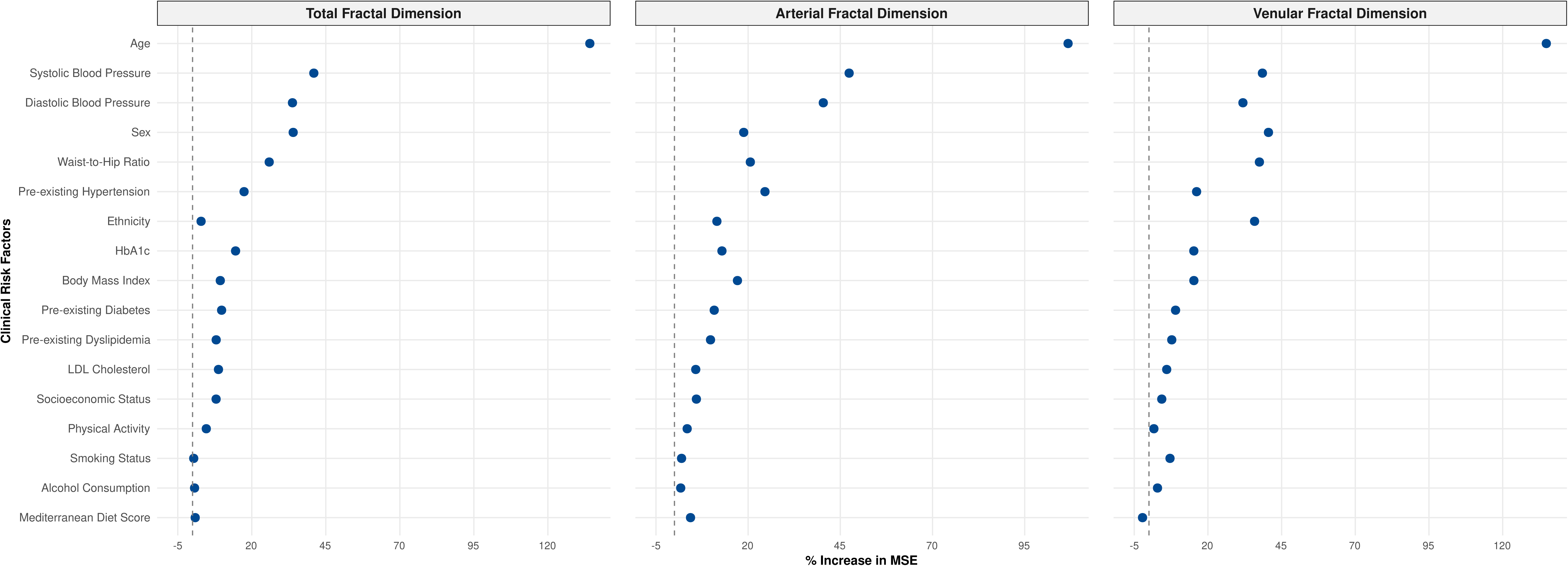
Determinants of total, arterial and venular retinal D_f_ based on random forest models. The x axis represents the mean decrease in accuracy for each clinical risk factor, representing its relative importance in predicting D_f_. Higher values indicate greater impact on model performance when the variable is permuted.

### Associations Between Retinal D_f_ and Incident CVD

Of the 4,781 participants free of CVD at baseline with good-quality retinal images, complete clinical data, and plasma metabolomic data, 546 (11.42%) developed CVD during follow-up (Supplementary Figure 3). The median follow-up duration was 5.75 years (IQR 5.48-6.04). Baseline characteristics of this sample are summarized in Table 1. Missingness across all covariates was low (Supplementary Table 4).

**Table 1.** Baseline characteristics of the sample stratified by incident cardiovascular disease (CVD) status.

|  |  | **No Incident CVD**<br>**(n = 4,235)** | **Incident CVD**<br>**(n = 546)** |
| --- | --- | --- | --- |
| Age, years, mean (SD) |  | 58.21 (8.14) | 62.92 (9.13) |
| Sex, male, n (%) |  | 1986 (46.89) | 284 (52.01) |
| Ethnicity, European, n (%) |  | 3934 (92.89) | 520 (95.24) |
| Body mass index, kg/m<sup>2</sup>,<br>mean (SD) |  | 27.72 (5.24) | 29.17 (6.08) |
| Systolic blood pressure,<br>mmHg, mean (SD) |  | 118.65 (15.41) | 124.14 (17.44) |
| Diastolic blood pressure,<br>mmHg, mean (SD) |  | 74.94 (9.49) | 75.45 (10.24) |
| Waist-Hip Ratio, mean<br>(SD) |  | 0.89 (0.10) | 0.92 (0.09) |
| HbA1c, nmol/L, mean (SD) |  | 5.52 (0.63) | 5.66 (0.77) |
| Low-density lipoprotein<br>cholesterol, nmol/L, mean<br>(SD) |  | 3.03 (0.89) | 2.96 (0.99) |
| Smoking status, n (%) |  |  |  |
|  | Non-Smoker | 2213 (52.26) | 241 (44.14) |
|  | Former Smoker | 1666 (39.34) | 250 (45.79) |
|  | Current Smoker | 356 (8.41) | 55 (10.07) |
| Frequency of alcohol<br>consumption, n (%) |  |  |  |
|  | Almost every day<br>(including 6 times<br>a week) | 572 (13.51) | 84 (15.38) |
|  | 4-5 times a week | 504 (11.90) | 61 (11.17) |
|  | 2-3 times a week | 1004 (23.71) | 108 (19.78) |
|  | Once a week | 530 (12.51) | 58 (10.62) |
|  | 2-3 times a month | 428 (10.11) | 56 (10.26) |
|  | About once a<br>month | 252 (5.95) | 29 (5.31) |
|  | Less than once a<br>month | 482 (11.38) | 70 (12.82) |
|  | Never | 394 (9.30) | 68 (12.45) |
| Physical activity, n (%) |  |  |  |
|  | Low | 1028 (24.27) | 167 (30.59) |
|  | Moderate | 2046 (48.31) | 254 (46.52) |
|  | High | 1161 (27.41) | 125 (22.89) |
| MDS score, n (%) |  |  |  |
|  | 0 | 10 (0.24) | < 6 |
|  | 1 | 47 (1.11) | 8 (1.47) |
|  | 2 | 231 (5.45) | 25 (4.58) |
|  | 3 | 437 (10.32) | 64 (11.72) |
|  | 4 | 710 (16.77) | 85 (15.57) |
|  | 5 | 904 (21.35) | 126 (23.08) |
|  | 6 | 847 (20.00) | 100 (18.32) |
|  | 7 | 606 (14.31) | 76 (13.92) |
|  | 8 | 315 (7.44) | 35 (6.41) |
|  | 9 | 84 (1.98) | 15 (2.75) |
|  | 10 | 13 (0.31) | < 6 |
| Socioeconomic status,<br>n (%) |  |  |  |
|  | Low | 261 (6.16) | 57 (10.44) |
|  | Moderate | 2113 (49.89) | 320 (58.61) |
|  | High | 1644 (38.82) | 144 (26.37) |
| Pre-existing dyslipidemia,<br>n (%) |  | 326 (7.70) | 60 (10.99) |
| Pre-existing diabetes,<br>n (%) |  | 537 (12.68) | 103 (18.86) |
| Pre-existing hypertension,<br>n (%) |  | 758 (17.90) | 165 (30.22) |

In unadjusted models, higher total, arterial and venular D_f_ values were significantly associated with reduced CVD risk (OR per 1-unit D_f_ increase: 0.62, 95% CI 0.53-0.72, p < 0.0001; OR: 0.78, 95% CI 0.70-0.86, p < 0.0001; OR: 0.74, 95% CI 0.67-0.82, p < 0.0001, respectively) (Table 2, models A). However, these associations were not statistically significant after adjustment for age and sex (Models B), or after further adjustment for the clinical risk factors (Models C) identified through forward stepwise selection: age, BMI, pre-existing hypertension, smoking status, WHR, physical activity, ethnicity, alcohol consumption, pre-existing dyslipidemia, and LDL cholesterol (Supplementary Figure 4). When D_f_ variables were modelled categorically, higher quartiles of total, arterial and venular D_f_ were associated with lower incident CVD in unadjusted analyses. For total D_f_, participants in third and fourth quartiles had 37% and 54% lower odds of incident CVD compared with the first quartile. Similar associations were observed for arterial D_f_ (26% and 47% decrease) and venular D_f_ (44% and 48% decrease) (Supplementary Table 5, Models A). However, these associations were not statistically significant after adjustment for age and sex (Models B) and fully adjusted multivariable models (Models C). Sex-stratified analyses revealed no evidence of effect modification by sex, regardless of whether D_f_ measures were modeled as continuous variables or quartiles. In the unadjusted ordinal logistic regression model, higher total, arterial and venular D_f_ was significantly associated with lower odds of being in a more severe CVD category, but these associations were not statistically significant after adjusting for covariates (Supplementary Table 6). Finally, when the composite CVD outcome was disaggregated, total, arterial and venular D_f_ were not independently associated with incident cardiovascular, cerebrovascular, or PVD events in the fully adjusted models. The only exception was arterial D_f_ in relation to cerebrovascular events, which remained marginally significant in the fully adjusted model (p = 0.02) (Supplementary Table 7).

**Table 2.**
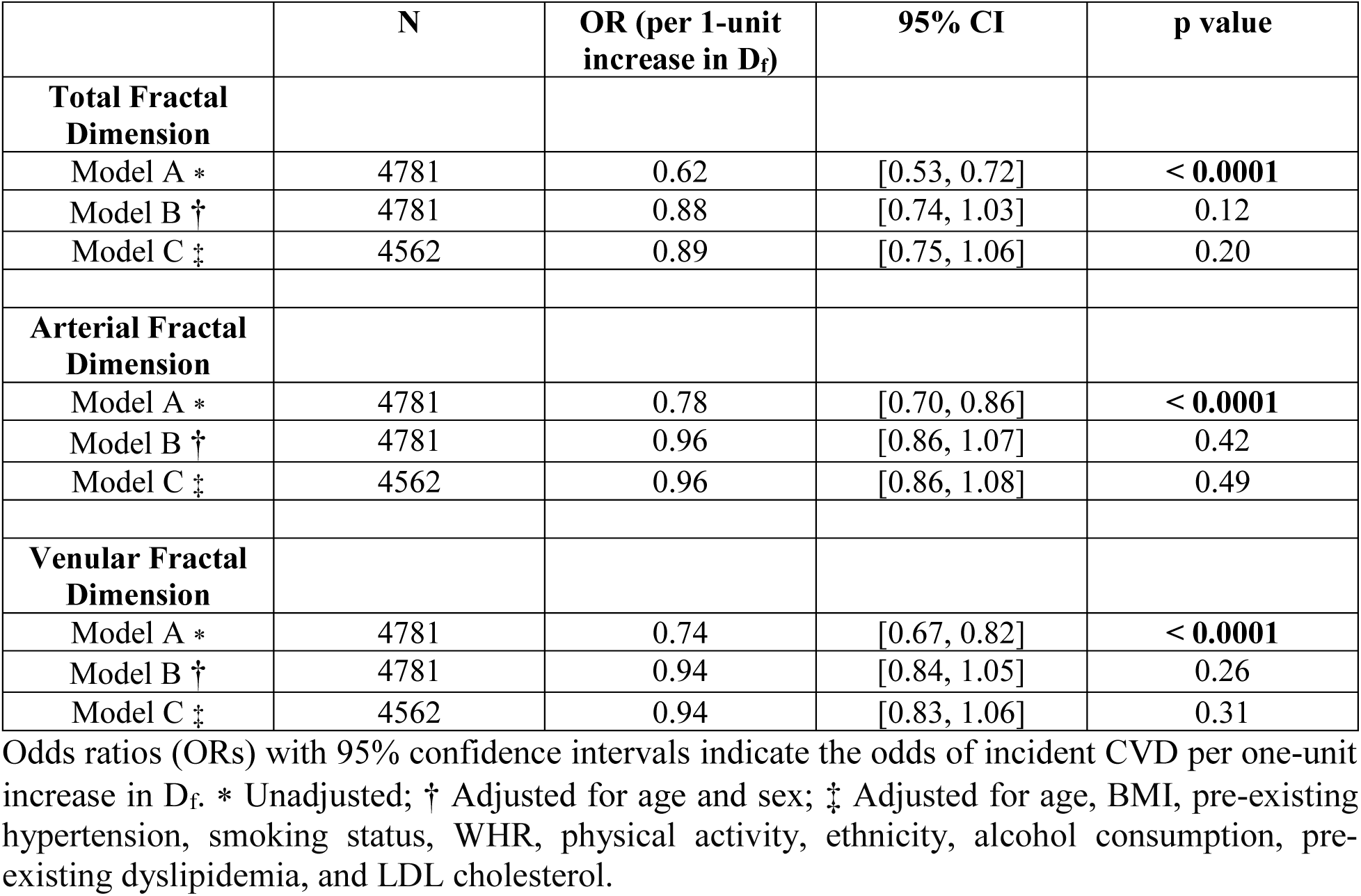
Associations between retinal D_f_ measures and odds of incident cardiovascular disease (CVD) in adults in the CLSA.

### Associations Between Retinal D_f_ Associated Plasma Metabolites and Incident CVD

We examined metabolome-wide associations between plasma metabolites and each of the three D_f_ measures, adjusted for age and sex. The results showed that 149 metabolites were associated with total D_f_, 147 with arterial D_f_ and 192 with venular D_f_ (all showing p < 0.05). After FDR correction, 43, 17, 85 metabolites remained significant for total, arterial and venular D_f_, respectively (q < 0.05) (Supplementary Tables 8-10). A total of 39 metabolites were associated with incident CVD in the fully adjusted model (p < 0.05) (Supplementary Table 11). Of these, 9 were also significant (p < 0.05) for one or more D_f_ measures (Table 3; Figure 2). After FDR correction, two widely known biomarkers of tobacco smoking, namely cotinine (HMDB0001046)^29^ and hydroxycotinine (HMDB0001390)^29^ remained statistically significant for both D_f_ and incident CVD (q < 0.05).

**Figure 2.**
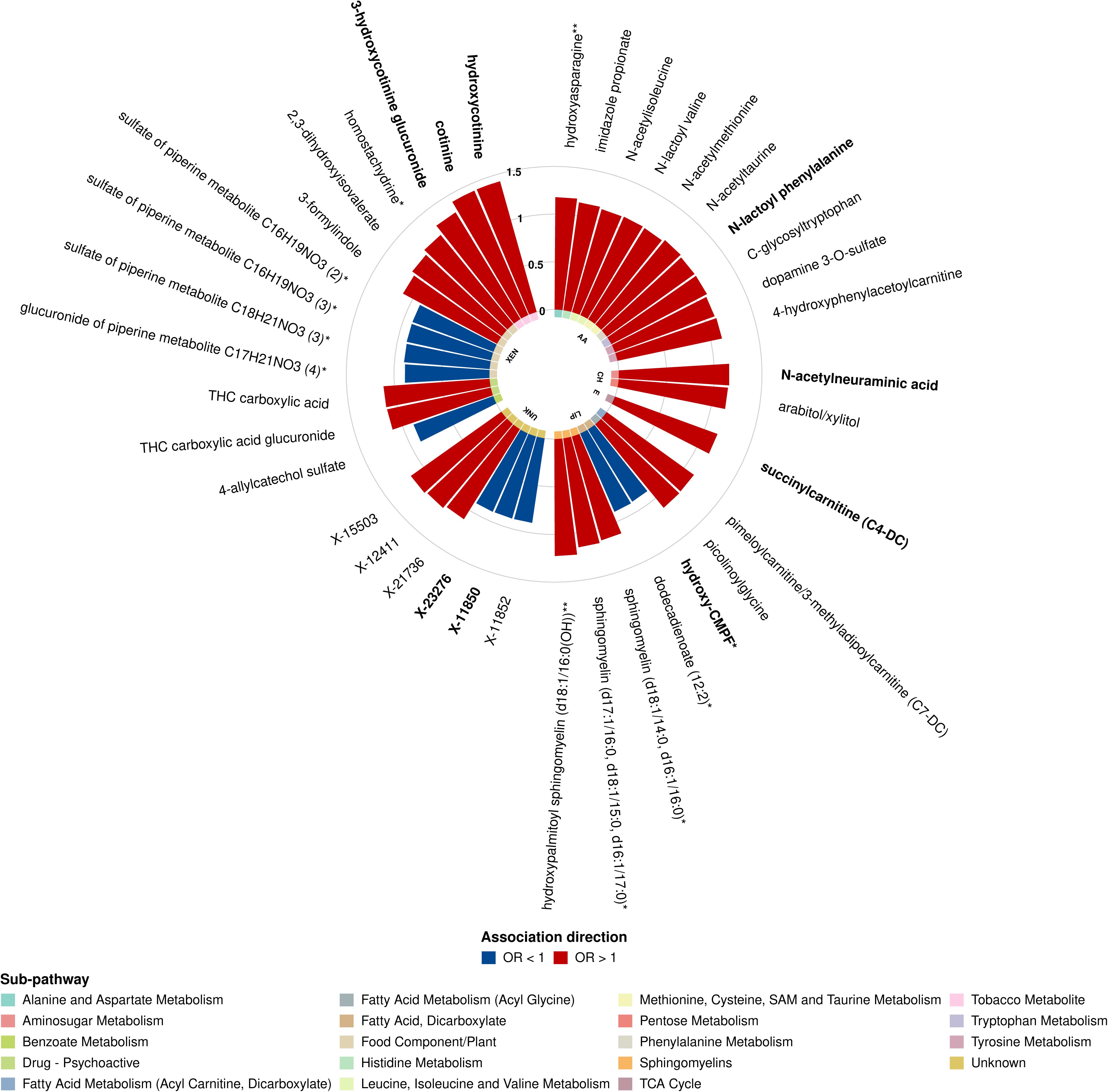
Circular plot of metabolites nominally associated with incident CVD. Metabolites are grouped by super and sub pathway. Odds ratios (ORs) were estimated using multivariable logistic regression, adjusted for age, BMI, pre-existing hypertension, smoking status, WHR, physical activity, ethnicity, alcohol consumption, pre-existing dyslipidemia, and LDL cholesterol. Blue and red bars indicate OR < 1 and OR > 1, respectively. Bolded metabolites were also nominally associated with retinal D_f_. Total-D_f_ associated metabolites include succinylcarnitine (C4-DC), hydroxy-CMPF, and X-23276. Arterial D_f_-associated metabolites include X-11850, cotinine, hydroxycotinine, and 3-hydroxycotinine glucuronide. Venular D_f_-associated metabolites include N-lactoyl phenylalanine, N-acetylneuraminic acid, and hydroxy-CMPF. Abbreviations: AA, amino acid; CH, carbohydrates; CV, cofactors & vitamins; E, energy; LIP, lipid; NT, nucleotide; UNK, unknown; XEN, xenobiotics.

**Table 3.** Summary of metabolites significantly associated with both retinal D_f_ and incident cardiovascular disease (CVD) (p < 0.05)

| Fractal Dimension | Superpathway | Subpathway | Metabolite Name | Linear Regression |  |  |  | Logistic Regression |  |  |  |
| --- | --- | --- | --- | --- | --- | --- | --- | --- | --- | --- | --- |
| | | | | $\beta$<br>(mean change in D<sub>f</sub> per 1 SD increase of metabolite) | SE | p value | q value | OR For CVD | 95% CI | p value | q value |
| **Total** | Energy | TCA Cycle | succinylcarnitine (C4-DC) | -0.02 | 0.01 | 0.03 | 0.20 | 1.14 | [1.03, 1.27] | 0.01 | 0.71 |
|  | Lipid | Fatty Acid, Dicarboxylate | hydroxy-CMPF* | 0.02 | 0.01 | 0.03 | 0.19 | 0.89 | [0.80, 0.99] | 0.03 | 0.77 |
|  | Unknown | Unknown | X-23276 | 0.02 | 0.01 | 0.01 | 0.13 | 0.90 | [0.82, 0.99] | 0.03 | 0.77 |
| **Arterial** | Unknown | Unknown | X-11850 | 0.03 | 0.01 | 0.02 | 0.17 | 0.90 | [0.81, 0.99] | 0.03 | 0.77 |
|  | Xenobiotics | Tobacco Metabolite | cotinine | -0.03 | 0.01 | 0.03 | 0.21 | 1.42 | [1.21, 1.66] | < 0.0001 | **0.01** |
|  | Xenobiotics | Tobacco Metabolite | hydroxycotinine | -0.03 | 0.01 | 0.03 | 0.21 | 1.42 | [1.20, 1.69] | < 0.0001 | **0.02** |
|  | Xenobiotics | Tobacco Metabolite | 3-hydroxycotinine glucuronide | -0.03 | 0.01 | 0.03 | 0.21 | 1.31 | [1.12, 1.53] | < 0.001 | 0.15 |
| **Venular** | Amino Acid | Phenylalanine Metabolism | N-lactoyl phenylalanine | -0.03 | 0.01 | 0.01 | 0.06 | 1.13 | [1.02, 1.25] | 0.02 | 0.77 |
|  | Carbohydrate | Aminosugar Metabolism | N-acetylneuraminic acid | -0.03 | 0.01 | 0.04 | 0.17 | 1.16 | [1.04, 1.29] | 0.01 | 0.67 |
|  | Lipid | Fatty Acid, Dicarboxylate | hydroxy-CMPF* | 0.03 | 0.01 | 0.02 | 0.13 | 0.89 | [0.80, 0.99] | 0.03 | 0.77 |

Linear regression analyses, adjusted for age and sex, show the associations between each D<sub>f</sub> measures and plasma metabolite levels. The beta coefficient represents the mean change in D<sub>f</sub> per 1 SD increase of metabolite, along with corresponding 95% confidence intervals and p values. Logistic regression analyses, fully adjusted for age, BMI, pre-existing hypertension, smoking status, WHR, physical activity, ethnicity, alcohol consumption, pre-existing dyslipidemia, and LDL cholesterol, show associations between D<sub>f</sub> related plasma metabolites and incident CVD. Odds ratios (ORs) with 95% confidence intervals indicate the odds of incident CVD per 1 SD increase in metabolite.

Notably, tobacco-related metabolites represented some of the strongest effect overall, with cotinine, hydroxycotinine, and 3-hydroxycotinine glucuronide (HMDB0001204)^29^ associated with lower arterial D_f_ and increased CVD risk. To capture the combined nicotine exposure from these metabolites, we computed a Total Nicotine Equivalent score (TNE-3), calculated by summing the raw ion responses for cotinine, hydroxycotinine, and 3-hydroxycotinine glucuronide prior to natural log transformation and standardization (Supplementary Figure 5), consistent with established approaches for quantifying total nicotine exposure^30^. Using this composite measure, TNE-3 was also associated with lower arterial D_f_ and increased CVD risk (Supplementary Figure 6).

Of the 4,781 participants, 2,454 were never smokers, 1,916 former smokers, and 411 current smokers. Nicotine metabolite levels differed significantly across smoking categories (Kruskal-Wallis p < 0.0001), with higher levels observed in current compared with former smokers (Wilcoxon p < 0.0001), as well as significant differences between former and never smokers (Wilcoxon p < 0.0001) (Supplementary Figure 7). These differences were reflected in a bimodal distribution of metabolite levels (Supplementary Figure 8).

Biochemical verification of smoking status using receiver operating curve-derived thresholds for cotinine, hydroxycotinine, and 3-hydroxycotinine glucuronide identified 530 metabolite-positive participants overall. Individuals were classified as metabolite-positive if levels exceeded the threshold for any one of the three plasma metabolites. Detection rates were highest among current smokers, with cotinine, hydroxycotinine, and 3-hydroxycotinine glucuronide detected in 93.7%, 92.9% and 82.5% of self-reported current smokers respectively, compared to 5.8%, 4.5%, and 3.0% among former smokers and 1.0%, 0.5%, and 0.4% among never smokers. Among the 530 biochemically verified smokers, all three metabolites were co-detected in 74.7% (n = 396), exactly two in 15.7% (n = 83), and exactly one in 9.6% (n = 51), with cotinine being the single most detected plasma metabolite (n = 41). Consistently, individuals classified as metabolite-positive had a nearly three-fold higher risk of incident lung disease (defined as physician-diagnosed emphysema, chronic bronchitis, chronic obstructive pulmonary disease, or chronic changes in lungs due to smoking) compared with metabolite-negative individuals (fully adjusted OR 2.83, 95% CI 1.47-5.15, p < 0.01).

Overall concordance between self-reported and metabolite-positive smoking status was high, with 385 participants (195 males, 190 females) classified as smokers and 4,225 (1,976 males, 2,249 females) classified as non-smokers in agreement across both measures. Concordance among non-smokers differed significantly by sex (Chi-square p value = 0.01). Discordant classification was observed in 171 participants including 145 individuals (81 males, 64 females) with detectable nicotine metabolites despite reporting no current smoking, and 26 individuals (18 males, 8 females) who reported current smoking without detectable metabolites; this also differed significantly by sex (Chi-square p value = 0.04).

Among the 145 metabolite-positive individuals who self-reported as non-smokers, 114 (60 males, 54 females) were former smokers and 31 (21 males, 10 females) were never smokers. Further characterization indicated that 12 participants (8.3%) reported occasional tobacco use, 19 (13.1%) reported use of nicotine replacement products, and 25 (17.2%) reported frequent passive smoke exposure (≥ weekly), while the remaining cases likely reflect residual exposure from prior smoking, low-level environmental exposure, or differences in timing between exposure and self-report.

Correlation analysis demonstrated that these plasma metabolites were correlated with each other (Spearman ρ = 0.69–0.89) and with self-reported smoking status (Spearman ρ = 0.41–0.52) (Figure 3). TNE-3 was strongly correlated with each individual metabolite (Spearman ρ = 0.69-0.99), with the strongest correlation observed between TNE-3 and cotinine and moderately correlated with self-reported smoking status (Spearman ρ = 0.41). Among current smokers, metabolite levels increased with self-reported smoking intensity (Supplementary Figure 9), with moderate positive correlations for cotinine (ρ = 0.43), hydroxycotinine (ρ = 0.40), 3-hydroxycotinine glucuronide (ρ = 0.34), and TNE-3 (ρ = 0.41) indicating that these biomarkers capture both smoking status and smoking intensity^31^.

**Figure 3.**
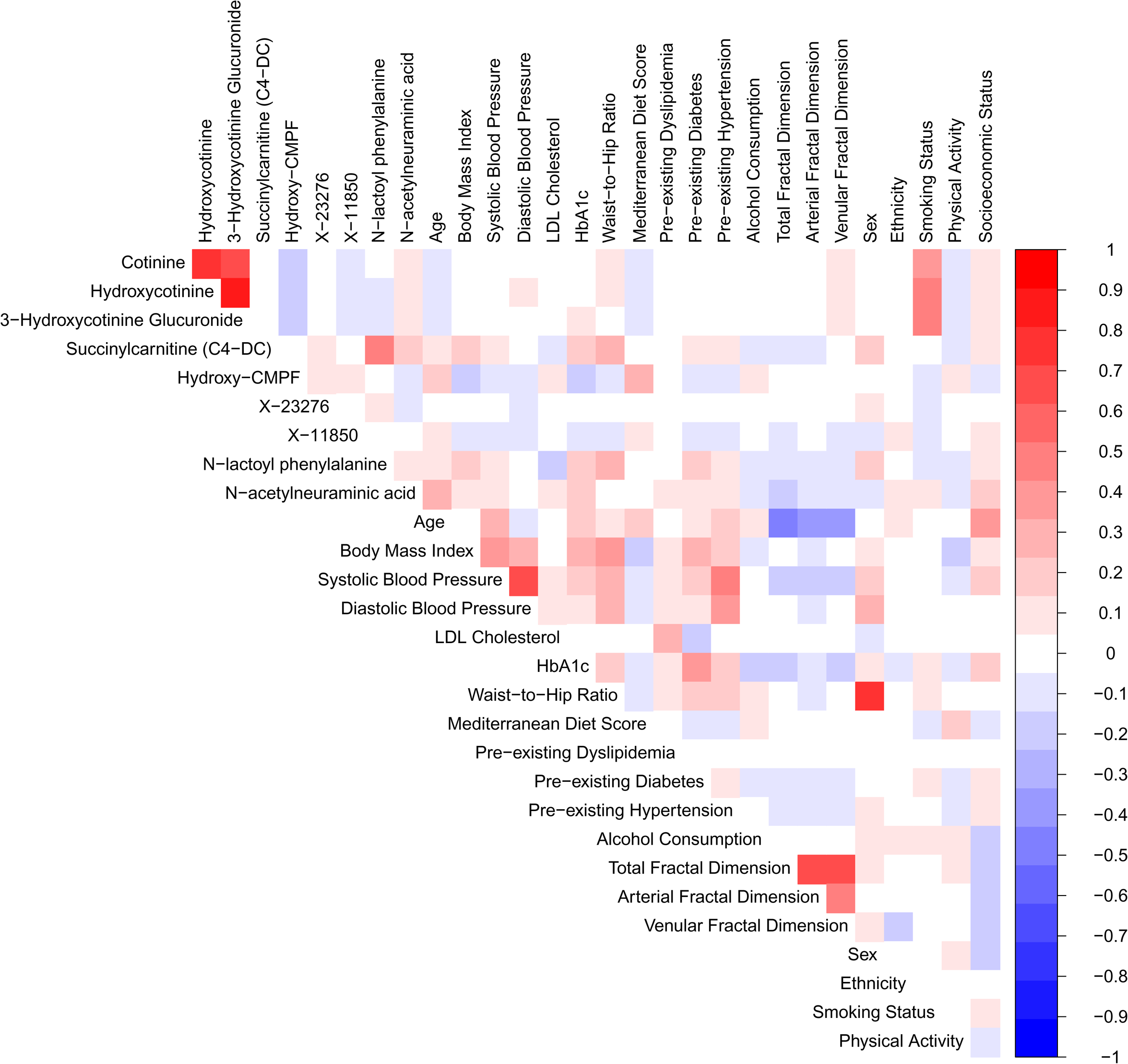
Correlation matrix of plasma metabolites, traditional cardiovascular risk factors, and retinal D_f_ measures. Heatmap showing pairwise Spearman correlation coefficients between circulating plasma metabolites, established cardiovascular risk factors and retinal D_f_ measures (total, arterial and venular). Color intensity represents the strength and direction of the correlations, with positive correlations in red and negative correlations in blue. Correlation coefficients were calculated using Spearman’s method.

To further assess interindividual variation in nicotine metabolism, we calculated the nicotine metabolic ratio (NMR), defined as the ratio of hydroxycotinine to cotinine^32^ (Supplementary Figure 10). Among biochemically verified smokers (n = 530), NMR was analyzed both as a continuous variable and categorically by quartiles, with Q4 representing fast nicotine metabolizers, and Q1-Q3 representing slower metabolizers (Supplementary Table 12). NMR was not significantly associated with incident CVD in any model, including analyses treating quartiles as ordinal categories and binary comparisons of Q4 versus Q1-Q3.

Beyond the nicotine metabolites, several additional compounds were associated with both retinal vascular complexity and CVD risk. Higher circulating levels of succinylcarnitine (HMDB0061717)^29^, part of the tricarboxylic acid cycle^33^, were associated with lower total D_f_ and increased CVD risk. Similarly, higher levels of N-lactoyl phenylalanine (HMDB0062175)^29^, shown to be involved in the regulation of appetite and energy expenditure^34^, and N-acetylneuraminic acid (or sialic acid) (HMDB0000230)^29^, part of the aminosugar metabolism pathway, were associated with lower venular D_f_ and increased CVD risk.

In contrast, several plasma metabolites were nominally associated with higher retinal vascular complexity and lower CVD risk. Hydroxy-3-carboxy-4-methyl-5-propyl-2-furanpropanoate (CMPF), part of the dicarboxylate pathway linked to fish and seafood intake within the Mediterranean diet^35^, was associated with higher total and venular D_f_ and linked to reduced CVD risk. Furthermore, two unidentified metabolites (X-23276, previously identified in the context of a carbohydrate dietary intervention^36^, and X-11850, predicted to be a tryptophan-related metabolite^37^), detected in reverse-phase negative ionization mode, were nominally associated with higher total and arterial D_f_ respectively and with a lower risk of incident CVD.

Furthermore, we examined associations of these nine D_f_-related plasma metabolites, including TNE-3, with cardiovascular, cerebrovascular, and PVD outcomes separately (Supplementary Table 13). For cardiovascular events, hydroxy-CMPF, cotinine, hydroxycotinine, TNE-3, and N-lactoyl phenylalanine demonstrated significant associations after full covariate adjustment (p < 0.05) that remained significant after FDR correction (q < 0.05). Cerebrovascular events showed fewer significant associations overall; cotinine, hydroxycotinine, 3-hydroxycotinine glucuronide, TNE-3, and X-23276 were significant after full adjustment (p < 0.05) but did not satisfy FDR correction, with X-23276 showing a significant inverse association specific to this outcome. For PVD, N-acetylneuraminic acid was significant after full adjustment (p < 0.05) but not after FDR correction. In contrast, hydroxycotinine, 3-hydroxycotinine glucuronide, and TNE-3 were significantly associated with PVD after FDR correction (q < 0.05). *Mediation Analysis of Metabolite-CVD Associations Through Retinal D_f_*

Mediation analyses were performed to determine whether the associations between exogenously derived plasma metabolites linked to both D_f_ measures and incident CVD were mediated through their corresponding D_f_ measures (Supplementary Table 14). For arterial D_f_, significant natural direct effects and total effects were observed for cotinine, hydroxycotinine, and 3-hydroxycotinine glucuronide (p < 0.05). In contrast, the natural indirect effects were not significant, and the proportion of the total effect mediated through arterial D_f_ was minimal (< 1%). A similar pattern was observed for TNE-3, which exhibited significant natural direct and total effects, but minimal mediation through arterial D_f_, with only 0.08% of the total effect attributable to the indirect pathway.

## DISCUSSION

Our study has shown that lower retinal D_f_ was associated with increased risk of incident CVD in unadjusted analyses. Total D_f_ showed the strongest overall predictive value for incident CVD compared to arterial and venular D_f_, but not independently of established CVD risk factors. Plasma metabolomics identified several metabolites linked to both retinal vascular complexity and CVD risk, with nicotine metabolites, particularly cotinine and hydroxycotinine, as well as the composite measure TNE-3, emerging as the strongest biomarkers.

Multiple studies have investigated the association between retinal D_f_ and incident CVD, with most reporting that lower D_f_ is associated with increased CVD risk^6^. However, these findings arise from studies that differ substantially in overall study design, outcome definitions^38^, approaches to quantifying retinal vascular complexity^39^, and population size and structure. Collectively, methodological differences, including the use of ICD-9 and ICD-10 codes for disease ascertainment, and variation in AI-based software used to quantify retinal D_f_ (e.g., the Retina-based Microvascular Health Assessment System (RMHAS) and Singapore I Vessel Assessment), may contribute to inconsistencies across studies and help explain the non-significant associations between D_f_ and incident CVD observed after multivariable adjustment in our cohort.

Furthermore, among the retinal D_f_ measures examined, total D_f_ demonstrated the strongest overall predictive value for incident CVD, likely as it captures overall microvascular complexity. Nevertheless, in this cohort total D_f_ did not provide prognostic information independent of established CVD risk factors, suggesting that it largely reflects age-related vascular changes already captured by clinical predictors^4^. Consistent with this, age emerged as the primary determinant of total, arterial, and venular D_f_. While total D_f_ captured the strongest overall signal, vessel-specific measures remain an important area for future investigation^40^. Discordance between arterial and venular D_f_ may help identify individuals with vessel type specific vulnerability that is not captured by total D_f_ alone.

Across the plasma metabolites examined, the top findings involved biomarkers of tobacco exposure. Cotinine and hydroxycotinine remained significantly associated with incident CVD after FDR correction, and together with 3-hydroxycotinine glucuronide were associated with lower arterial retinal D_f_. Consistent with these findings, their composite measure (TNE-3) was also associated with lower arterial D_f_ and increased CVD risk after FDR correction. Importantly, metabolite-defined smoking was strongly associated with incident lung disease, further reinforcing these biomarkers as robust indicators of tobacco-related systemic disease burden across both cardiovascular and pulmonary outcomes.

Given that plasma metabolite samples were collected at baseline between 2011 and 2015, prior to the widespread adoption of e-cigarettes^41^, elevated nicotine metabolite levels in this cohort most likely reflect exposure to conventional tobacco smoke from cigarettes. It is important to note that plasma cotinine and hydroxycotinine primarily reflect recent nicotine exposure over approximately the past 12 hours and therefore may not fully capture habitual smoking behaviour in all participants^42^. Beyond their role as exposure biomarkers, cotinine, hydroxycotinine, and 3-hydroxycotinine glucuronide are themselves tobacco-derived carcinogens, and elevated circulating levels have been associated with increased risk of lung and other smoking-related cancers^43^, further underscoring the systemic health burden of tobacco exposure captured by these plasma metabolites. These findings are biologically plausible as cigarette smoke generates reactive oxygen species (ROS), free radicals, and other pro-oxidants^44^, inducing chronic oxidative stress that damages the microvasculature and contributes substantially to CVD. However, mediation analysis suggests that arterial D_f_ explains only a small proportion of plasma metabolite-CVD associations, suggesting that additional mechanisms, such as genetic susceptibility, and unmeasured lifestyle or dietary factors not captured by the MDS we derived, contribute to risk.

These associations persisted even after adjustment for self-reported smoking status and were only modestly attenuated, indicating that these metabolites capture risk beyond reported smoking behaviour. Self-reported smoking status does not account for differences in nicotine intake, inhalation patterns, or individual metabolic variation, whereas circulating metabolites and the TNE-3 integrate both exposure and metabolic processing. Consequently, nicotine metabolites likely provide a more precise and biologically relevant measure of individual tobacco smoke exposure and metabolism.

The strong intercorrelation amongst cotinine, hydroxycotinine, and 3-hydroxycotinine glucuronide are consistent with their position within the nicotine metabolism pathway. Cotinine, the primary metabolite of nicotine, is converted in the liver by cytochrome P450 2A6 (CYP2A6) to hydroxycotinine, which is subsequently glucuronidated to 3-hydroxycotinine glucuronide via uridine 5’-diphospho-glucuronosyltransferases^45^. These sequential metabolic steps produce a tightly linked set of circulating biomarkers that reflect both nicotine exposure and individual metabolic capacity. The slightly weaker statistical signal observed for 3-hydroxycotinine glucuronide may reflect additional downstream metabolic variability and clearance processes that introduce greater biological variability in its circulating levels. However, the NMR, while an established phenotypic marker of CYP2A6 activity^32^, was not significantly associated with incident CVD across any adjustment model. The lack of association may reflect limited statistical power in this subgroup analysis, as biochemically confirmed smokers comprised a relatively small proportion of the overall sample, with modest event counts across quartiles reducing the ability to detect meaningful associations. In addition, cotinine and hydroxycotinine were primarily associated with cardiovascular events, while 3-hydroxycotinine glucuronide was more strongly linked to PVD and cerebrovascular outcomes, with TNE-3 showing associations across all three outcomes. To date, elevated plasma cotinine and hydroxycotinine levels have been linked to increased CVD risk^46^, consistent with evidence that cigarette smoke increases blood coagulability, a major risk factor for acute cardiovascular events^47^. Evidence for 3-hydroxycotinine glucuronide is limited, likely due to its less frequent measurement.

Beyond these findings, succinylcarnitine, hydroxy-CMPF, N-lactosyl phenylalanine, N-acetylneuraminic acid, X-11850, and X23276 showed suggestive associations with retinal D_f_ and incident CVD. However, most did not remain significant after correction for multiple testing, and their biological relevance to retinal microvascular structure remains uncertain. These exploratory signals may reflect broader metabolic pathways related to energy metabolism, inflammation, or diet, but further studies will be required to validate these associations and clarify their mechanistic relevance.

## Strengths and Limitations

A major strength of this study is the use of a large and well-characterized cohort, with metabolomics data available for nearly 10,000 participants. This is also one of few population-based cohorts globally to have retinal images available for > 10,000 individuals^48^. The CLSA’s broad baseline age range (45-85 years) captures mid-life exposures relevant to later-life health, offering an advantage over studies such as the Baltimore Longitudinal Study on Aging, which enrolled participants aged 65+^49^, and Atherosclerosis Risk in Communities Study which only enrolled participants aged 45-65^50^. Additionally, other U.S. longitudinal aging cohorts such as the Health and Retirement Study, Midlife in the United States, and Cardiovascular Health Study^50^, lack the integrated collection of retinal imaging and large-scale metabolomics data. In contrast, the CLSA allows for a comprehensive investigation of the relationships between retinal vascular complexity, circulating plasma metabolites and incident CVD across mid and later life.

However, several limitations should be noted. First, due to the absence of reliable time-to-event information for incident CVD, associations were assessed using logistic regression rather than Cox proportional hazards models, which may reduce sensitivity to detect associations compared with survival analyses. The cohort is predominantly of European ancestry, which may restrict the generalizability of findings to more diverse populations. Retinal D_f_ and circulating plasma metabolites were measured at a single baseline time point, limiting the ability to evaluate longitudinal changes and potentially underrepresenting habitual exposures. In addition, plasma metabolites reflect metabolic activity arising from multiple tissues and organs and do not provide tissue-specific information or distinguish between altered production and clearance, limiting mechanistic interpretation. The exclusion of low-quality retinal images may have resulted in a healthier analytic sample, as individuals with ocular or cardiometabolic conditions affecting image quality were less likely to be included. In multivariable models, only the two nicotine metabolites cotinine and hydroxycotinine, as well as TNE-3 remained significant with incident CVD after FDR correction, and so the findings should be interpreted cautiously until independently replicated.

Finally, the use of non-fasting plasma samples for metabolomics profiling may have introduced variability in the metabolite measurements.

In conclusion, retinal D_f_, which reflects microvascular complexity, is a predictor of incident CVD events, but not independently of established risk factors. Nicotine metabolites, particularly cotinine, hydroxycotinine, and TNE-3 emerged as the strongest biomarkers linking microvascular changes to CVD risk. Measuring these circulating metabolites, especially into composite scores, add predictive value over self-reported smoking status.

## Data Availability

Individual data used in this study can be provided by the Canadian Longitudinal Study on Aging (www.clsa-elcv.ca) pending scientific review and a completed material transfer agreement (requests for these data should be submitted to the CLSA).

## Acknowledgements

This research was made possible using the data/biospecimens collected by the Canadian Longitudinal Study on Aging (CLSA). This research was made possible using the data/biospecimens collected by the Canadian Longitudinal Study on Aging (CLSA). The development, testing and validation of the Short Diet Questionnaire (SDQ) were carried out among NuAge study participants as part of the Canadian Longitudinal Study on Aging (CLSA) Phase II validation studies, CIHR 2006-2008. The NuAge study was supported by the Canadian Institutes for Health Research (CIHR), Grant number MOP-62842, and the Quebec Network for Research on Aging, a network funded by the Fonds de Recherche du Québec-Santé. Funding for the Canadian Longitudinal Study on Aging (CLSA) is provided by the Government of Canada through the Canadian Institutes of Health Research (CIHR) under grant reference: LSA 94473 and the Canada Foundation for Innovation, as well as the following provinces, Newfoundland, Nova Scotia, Quebec, Ontario, Manitoba, Alberta, and British Columbia. This research has been conducted using the CLSA datasets [Comprehensive Baseline v7-2, Comprehensive Follow-up 1 v5-1, Comprehensive Follow-up 2 v4-1, Retinal Scans (Baseline), CLSA_QCNORMDATAALL_COMMONMETABOLITSONLY_12072024], under Application

Number 2304013. The CLSA is led by Drs. Parminder Raina, Christina Wolfson and Susan Kirkland. The authors gratefully acknowledge the time and commitment of the CLSA participants, without whom this research would not be possible. M.P. was supported by the E.J. Moran Campbell Internal Career Research Award from McMaster University and the Early Career Research Award from Hamilton Health Sciences (HHS). Retinal image analyses of the CLSA were supported by a New Investigator Fund from HHS (NIF-18453 to M.P.) and the CIHR, ACD-170312 to M.P. and PJT-178302 to M.P. P.B-M wished to acknowledge support from the Canada Foundation for Innovation and Genome Canada. The authors had full control, and the funders played no role in the design, analysis, or interpretation of this work.

## Disclaimer

The opinions expressed in this manuscript are the author’s own and do not reflect the views of the Canadian Longitudinal Study on Aging.

## Author Contributions

Writing - original draft: S.I, M.P. Conceptualization: M.P. Investigation: S.I, M.P. Writing - review and editing: P.B-M, T.R, D.J, E.T, F.M, S.M.A, M.C, M.P. Methodology: S.I, P.B-M, S.M.A, M.C, M.P. Resources: M.P. Funding acquisition: M.P. Data curation: S.I, M.P. Validation: M.P. Supervision: S.M.A, M.C, M.P. Formal Analysis: S.I, M.P. Software: S.I, E.T, M.P. Project administration: M.P. Visualization: M.P. All authors approved the final manuscript. The corresponding author attests that all listed authors meet authorship criteria and that no others meeting the criteria have been omitted.

## Supplementary Material

Tables S1 to S14 Figures S1 to S10

## Non-Standard Abbreviations and Acronyms

Df: Fractal dimension
VAMPIRE: Vessel Assessment and Measurement Platform for Images of the Retina
DCS: Data Collection Site
UPLC: Ultrahigh Performance Liquid Chromatography
CLSA: Canadian Longitudinal Study on Aging

